# Diverse Participant Recruitment for Infant Sequencing in the BabySeq Project

**DOI:** 10.1101/2024.10.01.24314717

**Authors:** Maya C. del Rosario, Sheyenne A. Walmsley, Barbara W. Harrison, Crystal T. Stephens, Bethany Zettler, Greysha Rivera-Cruz, Priyal Agrawal, Amy Brower, Stephanie Chigbu, Kurt D. Christensen, Casie A. Genetti, Richetta Givens, Nina B. Gold, Inez V. Reeves, Isabella Schichter, Habib Shariat, Sandra Simon, Hadley Stevens Smith, Melissa Uveges, Robert C. Green, Ingrid A. Holm, Stacey Pereira

## Abstract

**Purpose:** It is essential that studies of genomic sequencing (GS) in newborns and children include individuals from under-represented racial and ethnic groups (URG) to ensure future applications are equitably implemented. We conducted interviews with parents from URG to better understand their perspectives on GS research, develop strategies to reduce barriers to enrollment, and facilitate research participation.

**Methods:** Semi-structured interviews with 50 parents from URG.

**Results:** Nearly all parents (44) said they would be interested in participating in an infant GS study. Parents were interested in participating in GS research for reasons including clinical utility, personal utility, and/or family health benefits. Deterrents to enrollment cited by parents were discomfort with enrollment procedures (e.g., not wanting a heel stick), limited emotional bandwidth, unfavorable perceptions of the study, and concerns about potential results. Most parents (35 of 40) said they would want to receive all types of genetic results, including actionable and non-actionable, as well as childhood- and adult-onset.

**Conclusion:** Our findings demonstrate that parents from URG are interested in participating in GS research. Based upon these findings, we provide recommendations for designing GS studies that are responsive to their concerns.

## INTRODUCTION

When utilized as a screening tool for newborns and children, genomic sequencing (GS) has the potential to accelerate genetic diagnoses and to identify genetic risks and disorders before symptoms develop, allowing for the early initiation of preventive care and tailored therapy. Research studies that are inclusive of individuals from diverse populations and geographic locations, particularly under-represented racial and ethnic groups (URG) who have been historically excluded from genomics research, are critical to the equitable implementation of GS screening programs. However, historical research practices and ongoing medical racism have created lasting concerns among URG, including mistrust, privacy concerns, and fear of pain (1–4), resulting in lower rates of enrollment and participation. Understanding and addressing the concerns of parents from URG regarding genomic research participation will facilitate engagement of these communities in research and promote future implementation of GS screening programs in a way that benefit infants and families from all backgrounds.

The BabySeq Project is a randomized clinical trial (RCT) of GS in infants (1–4). In the first iteration of BabySeq we found that exome sequencing of newborns had clinical utility (2, 5, 6) and did not have negative psychosocial effects on families (7). However, although we offered enrollment to all families in large newborn nurseries and intensive care units that serve diverse populations, the generalizability of our results was limited because the families that chose to enroll were predominantly White, well-educated, and socioeconomically advantaged (8).

To address this lack of diversity, we initiated a new RCT of GS (using genome rather than exome sequencing), targeting enrollment to infants from pediatric primary care clinics that serve racially and ethnically diverse communities. Given the known concerns among URG communities, it is crucial to understand perspectives of individuals from these communities about participation in genomic research and develop strategies to overcome participation concerns and barriers. Prior to initiating enrolment in BabySeq, in order to inform the development of our study protocol, recruitment strategies, and disclosure methods, we conducted interviews with parents of children who received care from the BabySeq Project enrollment clinics, as well as a clinic that serves a similar population. Here we report interviewees’ hypothetical interest in, motivations for, and deterrents to participating in a study like the BabySeq Project; their preferences for learning about the study and being approached for enrollment; and their attitudes toward return of results and results sharing.

## MATERIALS AND METHODS

### Participants

Participants were parents of children who received their primary pediatric care at our future BabySeq Project enrollment sites: Children’s Hospital Primary Care Clinic (CHPCC) at Boston Children’s Hospital (BCH; Boston, MA), Mount Sinai Pediatric Associates Practice of Mount Sinai Hospital (MSH; New York, NY), and Children’s of Alabama Pediatric Primary Care Clinic at the University of Alabama Birmingham’s (UAB) Medical Center (Birmingham, AL). We also interviewed parents of children receiving care at Ambulatory Pediatrics at the Howard University College of Medicine (HUCOM; Washington, DC) due to its long-standing history of providing medical care to individuals from URG and successfully engaging those communities in research. Parents eligible to participate in the interviews were 18 years or older, had at least one child under 18 years of age, and spoke English or Spanish. Due to the project’s focus on enrollment of individuals from URG, we prioritized enrollment of parents of self-reported African American or Hispanic/Latino ancestry. The study was approved by the Institutional Review Boards (IRBs) at Boston Children’s Hospital and Howard University.

### Recruitment

Primary care providers (PCPs) of eligible parents briefly introduced the study and provided informational flyers in English or Spanish about the study during routine visits. Contact information for interested parents was forwarded to the study team. A member of the study team reached out to interested parents via email and phone to offer participation.

### Interviews

A semi-structured interview guide was developed by the study team and reviewed by the BabySeq Community Advisory Board (CAB) for content, language, clarity, and cultural sensitivity. The CAB was convened at the initiation of the second iteration of the BabySeq Project before enrollment began and comprises parents, community leaders, and previous BabySeq participants from the communities from which we are currently enrolling. A brief educational intervention with supporting PowerPoint slides (see Supplemental materials), which included a description of genes, genetic variants, standard newborn screening, and the goals of the BabySeq Project, was delivered prior to interview questions about parents’ perspectives on GS in infants. Demographic information was self-reported at the conclusion of the interview.

Interviews were conducted in English (MCD) or in Spanish by a native-Spanish speaking team member (GRC) by videoconferencing with Zoom, audio-recorded, and the audio files were transcribed following the removal of identifying information. Transcripts of interviews conducted in Spanish were professionally translated into English prior to analysis. Parents were compensated with a $50 gift card for their participation.

### Interview analysis

We used the online qualitative analysis software Dedoose (https://www.dedoose.com) to manage the coding process. A codebook of broad deductive codes was developed based on our research questions and the extant literature (e.g., deterrents to enrollment). Each interview was coded by three researchers (MCD, SAW, and BWH) using the codebook and then compared for a team-based coding approach. Discrepancies were discussed amongst researchers until consensus was reached for each coded segment. New inductive codes were added to the codebook iteratively, as needed, when predetermined codes did not match our data. We then used an inductive, two-step abstraction process to first identify themes within each broader deductive code, and then categorize the inductively identified themes together into larger, overarching themes(9). Each segment of text was reviewed by two of four research team members (MCD, SAW, IAH, SP) and all abstractions were discussed until consensus was reached.

## RESULTS

### Study Population

We interviewed 50 participants (Table 1) including 30 from HUCOM, 8 from BCH, 7 from MSH, and 5 from UAB. Five interviews were conducted in Spanish and the rest were in English (90%). Parents primarily self-identified as African American or Black (62%), non-Hispanic (64%), female (92%), were between 20-35 years of age (70%), and had some college education (52%). Most parents had more than one child (62%) and most had at least one child who was less than 5 years of age (98%). Most parents had no prior experience with genetic testing (60%). The majority of those who reported average household income indicated that it was below $50,000 (56%).

**Table 1.** Participant Characteristics.

|  | Participants, n (%)* |
| --- | --- |
| **Place of Enrollment** |  |
| Boston Children's Hospital | 8 (16) |
| Mount Sinai Hospital | 7 (14) |
| University of Alabama Hospital | 5 (10) |
| Howard University | 30 (60) |
| **Gender** |  |
| Female | 45 (92) |
| Male | 5 (8) |
| **Age (years)** |  |
| <30 | 23 (46) |
| 30-35 | 12 (24) |
| 36-40 | 4 (8) |
| 41-45 | 10 (20) |
| 46-50 | 1 (2) |
| **Number of Children** |  |
| 1 | 19 (38) |
| 2 | 11 (22) |
| 3 | 8 (16) |
| 4 | 5 (10) |
| 5 | 2 (4) |
| 6+ | 4 (8) |
| Unknown | 1(2) |
| **Self-reported Race** |  |
| Black, African-American, or African | 31 (62) |
| White | 15 (30) |
| Unknown | 4 (8) |
| **Self-reported Ethnicity** |  |
| Non-Hispanic | 32 (64) |
| Hispanic | 13 (26) |
| Unknown | 4 (8) |
| **Education Level** |  |
| Some High School | 5 (10) |
| High School Graduate or GED | 19 (38) |
| Some College | 8 (16) |
| Bachelor's Degree or Equivalent | 13 (26) |
| Master's Degree | 4 (8) |
| Unknown | 1 (2) |
| **Household Income (\$)** | |
| <15,000-24,999 | 13 (26) |
| 25,000-49,999 | 12 (24) |
| 50,000-99,999 | 5 (10) |
| >100,000-150,000 | 5 (10) |
| Unknown or declined to respond | 14 (28) |

\*N varies because not all data was collected for all interviews

### Results Overview

Nearly all parents (44) said they would be interested in participating in an infant GS study, noting several motivations including clinical utility, personal utility, and/or family health benefits. While only five parents said they would be unlikely to participate in GS research, many parents named reasons that might deter other parents from enrolling, including discomfort with enrollment procedures (e.g., not wanting a heel stick), limited emotional bandwidth, unfavorable perceptions of the study, and concerns about potential results they could receive. See Table 2 for themes of motivations and deterrents to study participation. When asked about results preferences, most parents (35 of 40) said they would want to receive all types of genetic results that could be available, including results for preventable conditions, treatable conditions, conditions with childhood onset, conditions with onset only in adulthood, and conditions that are not treatable or preventable.

**Table 2.**
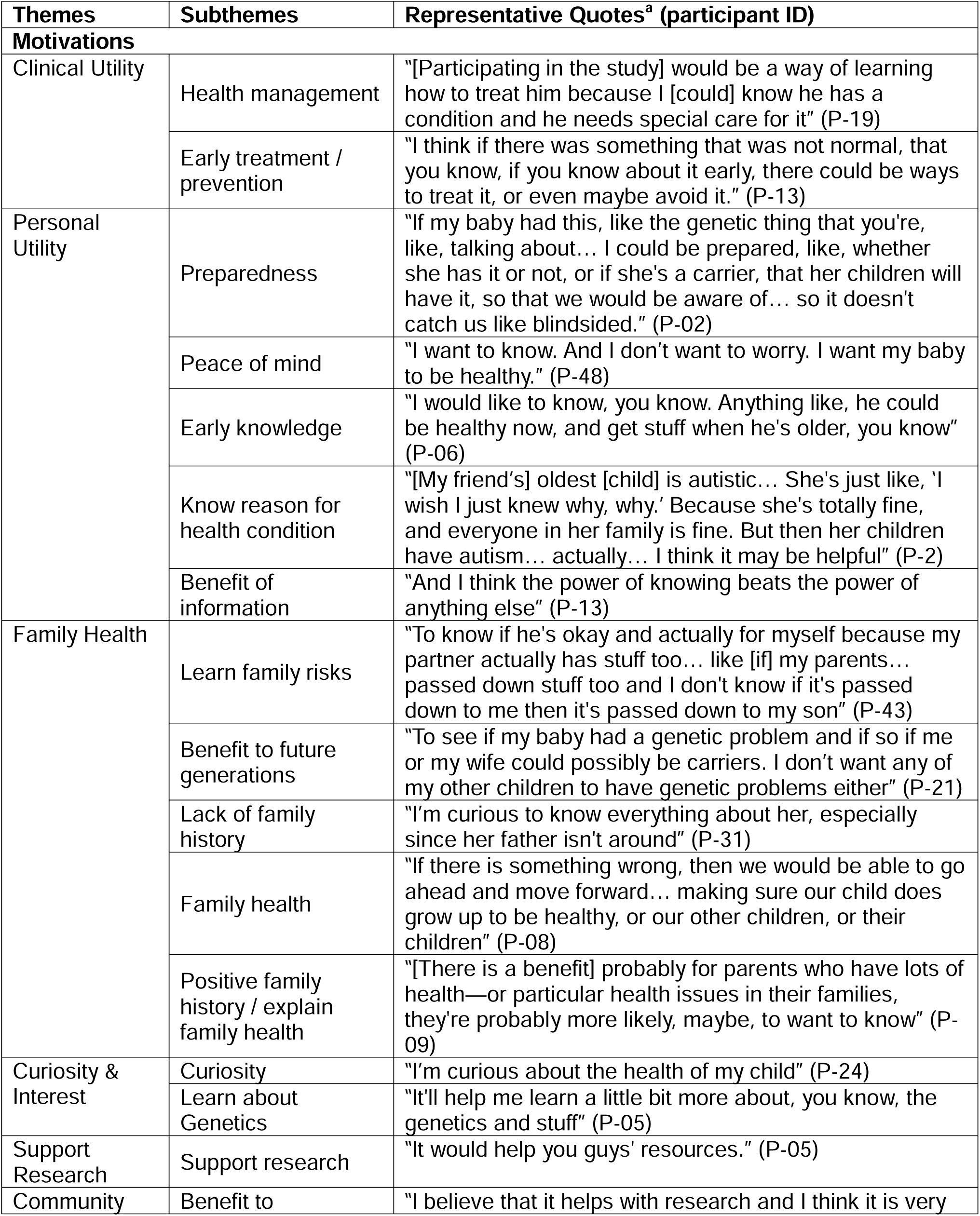
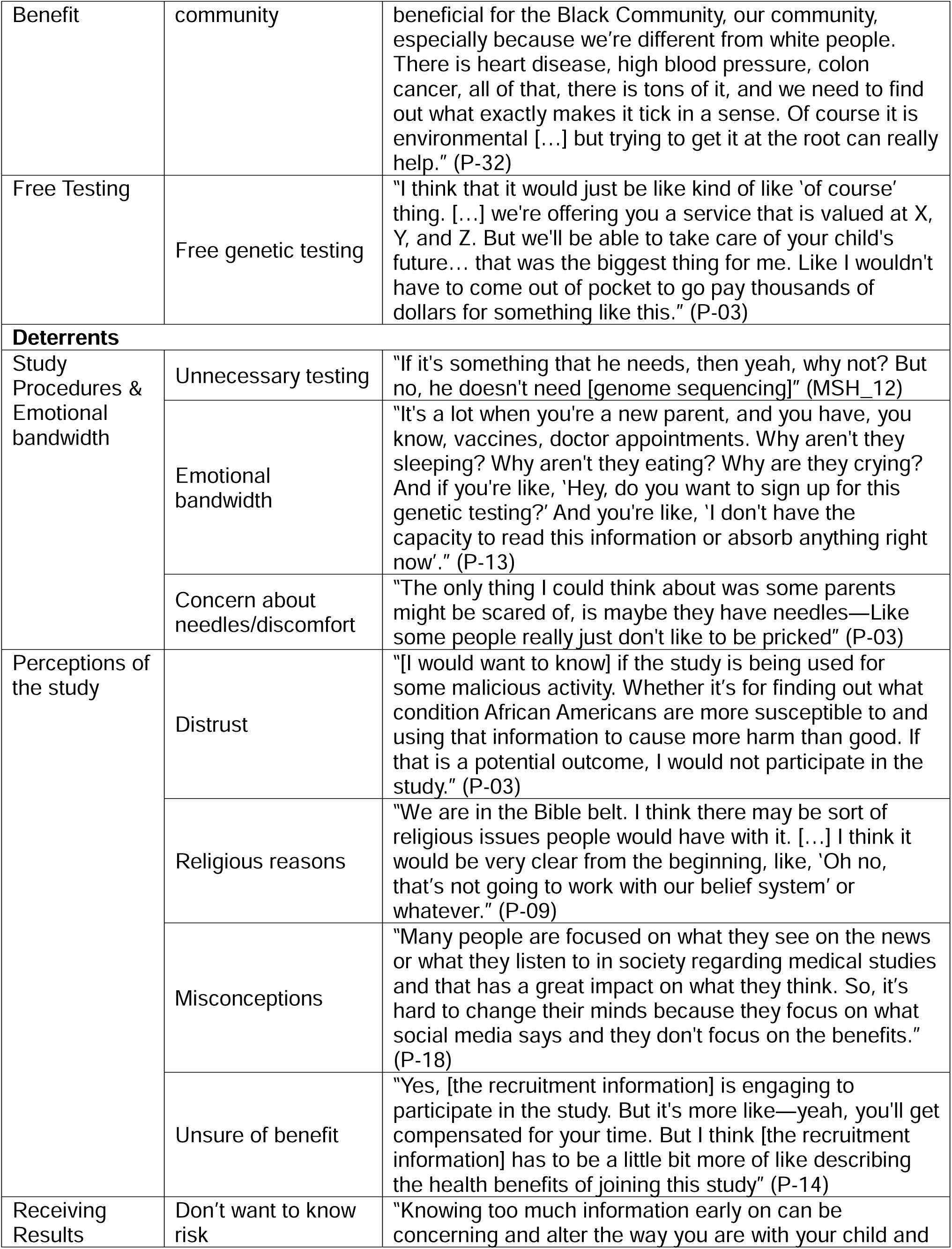
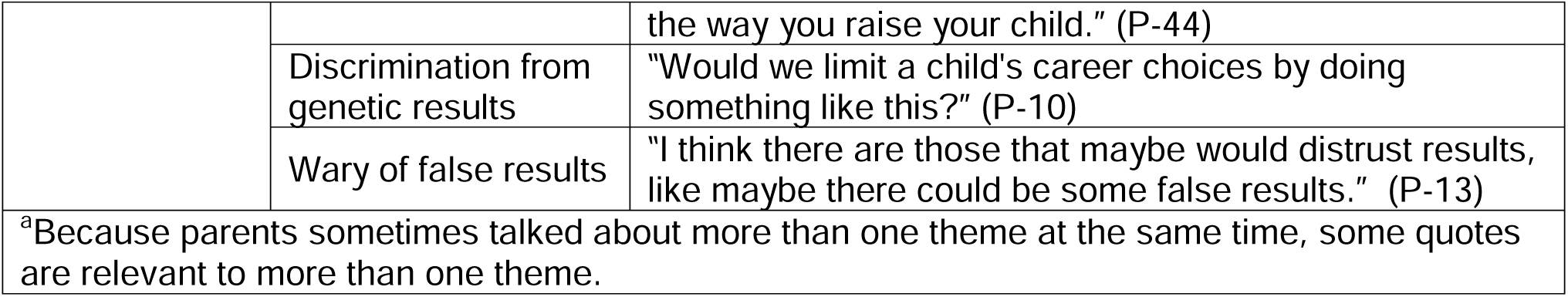
Domains, themes, and representative quotes for motivations and deterrents to study participation.

### Receiving information about the study

After we described an infant GS research study, we asked interviewees how they thought parents should be introduced to such a study. Most parents said the study should be introduced by a clinician that the parent knows, either the child’s PCP or another one of their baby’s clinicians. Other suggested methods for informing parents about the study included by letter, email, or phone call, or via flyers, brochures, or pamphlets at the clinical practice.

Parents were asked how they would like to get more information about the GS study before deciding whether to enroll. Most suggested that the study coordinators provide information using a study website, video, flyers, brochures, or pamphlets. Other parents suggested that study personnel speak directly to the parent’s own PCP or their child’s doctor, connect them with currently enrolled parents, or offer additional opportunities to speak with members of the research staff. Some parents offered suggestions about additional information that would help them decide about participating in the study, such as information about the types of diseases that could be screened and the timeline for their engagement in the study.

One parent noted the importance of keeping cultural sensitivity in mind when approaching eligible parents for enrollment, explaining that they felt it was important for study coordinators to understand the cultural background of the potential participants and to approach parents in ways that aligned with their cultural expectations.

### Motivations for enrollment

Most parents (44) said they would be very likely to enroll in an infant GS study like BabySeq. All parents were asked about reasons they might want to or not want to enroll. We identified seven overarching themes of motivations for enrolling (Table 2). One of the most cited reasons was the potential for clinical utility. Some parents specifically noted that early treatment or prevention of health conditions would be the reason they would choose to enroll. Others provided less specific reasoning but said they’d be interested in enrolling for purposes of managing their child’s health, such as knowing how to best care for their child.

Another common motivation parents noted was personal utility. Parents’ definition of personal utility included the general benefit of information, knowing a reason for a health condition, and early knowledge, all of which were described without being linked to specific clinical or health benefits. Parents also described being able to be prepared, or the potential for GS to provide peace of mind.

Many parents cited family health as a motivation for enrollment. Some parents noted the potential for GS to benefit the health of other members of the child’s family. Others noted specific family health benefits, including the possibility for GS information to be beneficial in cases where family health history was unknown, to explain known family history of conditions, or to learn about genetic risk in the family. One parent described how this could be beneficial for reproductive planning and could improve the health of future generations.

### Deterrents to enrollment

Five parents initially indicated that they would personally not be interested in joining an infant GS study. Among these five parents, four explained that they felt this testing was unnecessary given all the other routine screening children receive. Another parent said they would not want to enroll their child because GS utilized as screening in infants is not an established practice. One parent also said they were fearful about this kind of screening but did not explain why, and another expressed that they were unlikely to have enough time to commit to participating in a GS study.

Although the remainder of parents reported they would be likely to join an infant GS study, some offered reasons why they believed that other parents might not enroll. The most cited hypothetical deterrent for a parent not enrolling was not wanting to know the results of testing. Several parents speculated that some parents might be concerned that the results could negatively impact their quality of life, or that a genetic result suggesting a child was at risk for developing a health condition may change the way parents viewed their child.

Distrust and misconceptions about genomic research were mentioned by 5 parents as potentially deterring other parents from enrolling. Some parents indicated that other parents may not participate because they wouldn’t fully understand or trust the motives of the research team and would worry that the study could have malicious intent or plans to share data in ways that could harm parents or communities. Others described how the history of mistrust between URG and medical systems could dissuade some from enrolling in a GS study, specifically noting that “the Black community hasn’t had the best relationship [with doctors and healthcare institutions].” One parent suggested that misconceptions driven by media portrayal of medical research and negative social media may deter parents from enrolling.

Many parents expressed that logistical issues might deter them, or others, from participating in a GS study. These issues included not having enough available time to devote to the study (e.g., for appointments or surveys), inability to travel for study-related appointments, struggles with taking time off work to participate, and being too busy. Some parents were concerned that collecting a blood sample from their child would cause unnecessary pain or discomfort.

### Return of results

The majority of parents (35) expressed that they would be interested in receiving all types of results described, including results for preventable conditions, treatable conditions, conditions with childhood onset, conditions with onset only in adulthood, and conditions that are not treatable or preventable. When asked why they would like to receive such results, some parents expressed interest in knowing their child’s risk for family planning purposes, and others noted a desire to know their child’s genetic risk to be as prepared as possible to take care of their child as they get older.

When asked how they would like to receive their child’s results, most parents (41) said they would prefer to receive results in person, while 28 said that receiving results via video call would also be acceptable. Fewer parents (11) felt comfortable receiving results via phone or letter. When asked which types of materials they thought could facilitate better understanding, 27 parents were comfortable receiving the written report, 21 suggested adding pictures, graphics, or other visual aids, and 19 suggested a study website or video with additional information. When asked with whom they would share their child’s results, 39 parents said that they would share results with their child’s doctor and would want the results placed in their child’s medical record for all the clinicians involved in their child’s care to access.

Many parents gave recommendations for the study to provide care after results disclosure, including providing referrals to specialists and follow-up care, and more information about risk-reduction recommendations. Some parents also mentioned that being connected to support and advocacy groups would be personally useful.

### Sharing results with family members

A total of 42 of 47 parents said they would share their child’s genetic testing results with close family members and would suggest other family members pursue genetic testing, if recommended. Of those parents, some specified that who they would share their child’s results with would depend on both their biological relationship (e.g., first-degree relatives) and emotional closeness with their relative. Parents also mentioned barriers, including family dynamics and location of family members, that may prevent extended family from receiving testing. Almost all parents (47 of 48) indicated that if their child had a result that suggested their child was at risk for developing a health condition, it would be important to have themselves and their child’s other parent tested so they could understand their own risk and the risk for their future children. Only one parent said they would not want to proceed with testing themselves because they felt it would be too late to inform their health management.

## DISCUSSION

In the first iteration of The BabySeq Project (3) we demonstrated that newborn sequencing could identify clinically important health risks (2, 5, 6) in infants without causing psychosocial harm to families (7). However, like other studies on newborn sequencing to date (10), the first iteration of BabySeq lacked adequate enrollment of individuals from URG. To ensure future applications of GS in infants are equitably implemented, it is critical that we enroll URG participants in GS research. To do so, it is crucial to engage directly with members of URG to not only better understand and address concerns and barriers that may impede their enrollment, (11, 12) but also identify factors that may encourage and facilitate participation. In this study, our interviewees were largely positive toward hypothetically participating in a study of infant GS, but also identified potential deterrents to enrollment. They expressed preferences regarding various aspects of the study protocol that may not have otherwise been anticipated by our study team and provided suggestions to improve the implementation of GS research and future population-wide genomic screening programs.

Many of our findings in this interview study resonate with recurring themes in prior work assessing parents’ attitudes toward infant GS. Our previous research in the first iteration of BabySeq, and others (13), found that parents anticipated clinical and personal utility of newborn sequencing results, which aligns with the motivations for research participation identified in this study (14). In this study, we found that most parents expressed a preference to receive genomic results related to a wide range of health conditions, including adult-onset only conditions and conditions that are not treatable or preventable, the disclosure of which are currently controversial (15–17). These findings are consistent with our previous work (18) and other studies on preferences for genomic results both within and outside the context of infant GS (19, 20). Likewise, many of the deterrents to enrollment our participants identified reflect actual reasons parents declined the first iteration of the BabySeq Project, such as the additional burden the study could pose, concerns about what results might be received, and potential for discrimination (8). Interestingly, many of our interviewees did not view these concerns as barriers to their own enrollment in an infant GS study, but instead suggested that others may perceive these concerns as barriers.

Other potential deterrents identified by our interviewees were more closely aligned with established research about reasons for declining research among members of URG, including distrust of and potential for misconceptions about the study (21, 22). One strategy to foster trust that was emphasized by both interviewees and our CAB was to involve a clinician known to the family to introduce the study. This suggests the inclusion of clinicians who already have an established rapport with families considering participation can help address concerns. Other recommendations for designing infant GS studies that reduce barriers and facilitate enrollment among URG from our research findings cover topics such as sample collection methods, clinical follow-up, support to ease burden on participants, and efforts to reduce distrust and misconceptions; see Table 3.

**Table 3.**
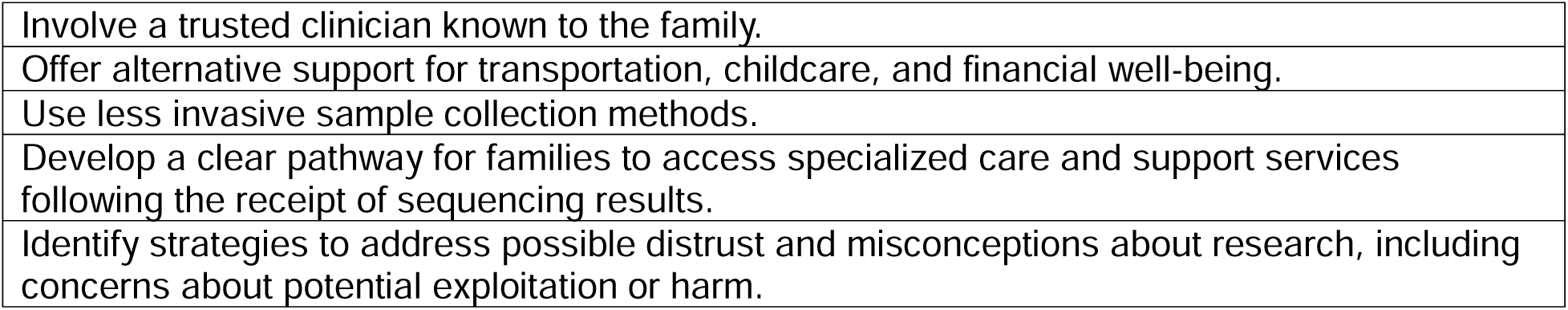
Recommendations to facilitate NBSeq research participation among URG.

Our results should be interpreted within the study’s limitations. This work was qualitative, and as such, the goal was not to generate generalizable data, but rather findings that can be transferable to similar contexts, such as other studies enrolling members of URG in newborn and childhood GS or similar research (23). Another limitation of our study is that the results we have reported are in response to hypothetical questions about interviewees’ interest in participating in an infant GS study; it is possible that they may have felt differently if approached to actually enroll in the study. Similarly, there is the possibility that parents may have been inclined to answer more favorably toward infant GS research in order to appease the interviewer (i.e., social desirability bias). Finally, because our interviewees were parents who agreed to participate in this interview study, it is possible our data reflect the attitudes of individuals who are more open to research participation in general, particularly regarding genomics. Indeed, nearly all our interviewees said they would be interested in participating in an infant GS study while also noting deterrents to participation that may cause other parents to choose not to enroll. Our previous work has demonstrated the potential for actual enrollment to be less than rates suggested in hypothetical situations (8, 24).

Our findings support that individuals in URG are interested in genetics research, and direct engagement with both the general population and URG to understand and address concerns and barriers that may impede the enrollment of diverse populations is critical. Earning trustworthiness with URG is essential to produce meaningful research outcomes that will allow diverse populations to confidently access the benefits of genomics. Our findings and recommendations can be used to help design studies of infant GS that are responsive to the concerns among URG that reduce barriers to enrollment.

## Data Availability

All data produced in the present study are available upon reasonable request to the authors

## Acknowledgements

We sincerely thank the members of the BabySeq Community Advisory Board for their valuable contributions in reviewing the semi-structured interview guide. Their insights were instrumental in refining the tool to ensure its relevance and cultural sensitivity. Special thanks to Crystal T. Stephens, Richetta Givens, Angela G. Williams, Roxann C. Harvey, Delante Lee Bess, Gabriela Bess, Trinisha Williams, Sarita Edwards, Erica Y. Lopez, and Alyssa R. Carter for their time, thoughtful feedback, and commitment to this project.

## Author Contributions

Conceptualization: MCD, SAW, BWH, BZ, CAG, IAH, SP; Data Curation: MCD, BWH, IS, SAW, IAH, SP; Formal Analysis: MCD, SAW, BWH, BZ, IS, CTS, IAH, SP; Funding Acquisition: IAH, RCG; Investigation: MCD, SAW, BWH, CTS, BZ, GRC, SC, SS, IAH, SP; Methodology: MCD, SAW, BWH, BZ, CTS, GRC, IAH, SP; Project Administration: MCD, SAW, IAH, SP; Supervision: IAH, SP; Writing – original draft: MCD, SAW, BWH, CTS, IAH, SP; Writing – review & editing: MCD, SAW, BWH, CTS, BZ, GRC, PA, AB, SC, KDC, CAG, RG, NBG, IVR, IS, HS, SS, HSS, MKU, RCG, IAH, SP.

## Ethics Declaration

The study was approved by the Institutional Review Boards (IRBs) at Boston Children’s Hospital and Howard University. Informed consent was obtained from all participants.

